# Joint Modeling of Hypertension, Diabetes, and Cardiovascular Disease in Uganda: A Gaussian Copula Approach

**DOI:** 10.1101/2025.07.02.25330705

**Authors:** Grace Kakaire, Edna Chumoh

## Abstract

Non-communicable diseases (NCDs), including hypertension, diabetes mellitus, and cardiovascular disease (CVD), are increasingly responsible for morbidity and mortality in low- and middle-income countries (LMICs). In Uganda, these conditions are becoming more prevalent due to rapid urbanization, lifestyle transitions, and demographic shifts. Despite their frequent co-occurrence, existing research typically investigates each condition in isolation, failing to capture their shared etiology and potential dependence.

This study analyzes nationally representative data from the 2014 Uganda WHO STEPS survey to assess both individual-level risk factors and the joint distribution of hypertension, diabetes, and CVD.

A total of 3,987 adults aged 18–69 years were included. Survey-weighted logistic regression models were fitted for each binary outcome, adjusting for age, sex, region, marital status, body mass index (BMI), education, income, physical activity, cholesterol, tobacco, and alcohol use. Missing data were addressed using multiple imputation by chained equations. Probability-scale residuals from the marginal models were used to fit both Gaussian and Clayton tri-variate copulas, enabling flexible modeling of the dependence structure.

Hypertension affected 62.3% of participants, diabetes 65.8%, and CVD 9.3%. Obesity was strongly associated with hypertension (OR = 2.00; 95% CI: 1.02–3.92), while overweight and obesity were inversely associated with diabetes. Age was positively associated with both hypertension and CVD. Women had lower odds of hypertension but higher odds of CVD. Participants in the Eastern region had lower CVD risk, while those in the Northern region showed elevated hypertension risk. The Gaussian copula showed a better fit than the Clayton copula (log-likelihood: 184.04 vs. 87.33; AIC: –362.08 vs. – 172.65), indicating moderate symmetric dependence between the outcomes. The Clayton copula revealed minimal lower-tail dependence (Kendall’s τ = 0.0365), suggesting weak clustering of low- outcome probabilities.

These findings highlight the need for integrated, person-centered approaches to NCD care in Uganda. Joint modeling using copula-based frameworks enhances understanding of multi-morbidity patterns and enables better targeting of prevention and intervention strategies. The Gaussian copula’s superior performance underscores the potential of symmetric dependence models in capturing the clustering of NCDs, particularly the strong correlation observed between hypertension and CVD. These insights are essential for informing health policy and optimizing resource allocation in settings undergoing epidemiological transition.

## Introduction

Non-communicable diseases (NCDs) such as hypertension, diabetes mellitus, and cardiovascular disease (CVD) have emerged as leading causes of morbidity and mortality globally, accounting for over 70% of all deaths (World Health Organization [1, 2]. While traditionally considered a concern for high- income countries, recent decades have witnessed a dramatic epidemiological shift, with low- and middle- income countries (LMICs) increasingly bearing the brunt of the NCD burden [3, 4]. In sub-Saharan Africa, this shift has been accelerated by rapid urbanization, changes in dietary habits, sedentary lifestyles, and increased life expectancy [5]. As a result, the region is now grappling with a rising tide of chronic illnesses amidst enduring infectious disease challenges and constrained health systems [6].

Uganda exemplifies this trend, with national health surveys and facility-based data indicating a rising prevalence of NCDs. Hypertension affects nearly one in four adults, and diabetes prevalence is steadily increasing, particularly in urban centers [7–9]. Cardiovascular disease, while less commonly diagnosed, contributes significantly to premature mortality and disability, often going undetected until advanced stages [10]. These conditions frequently share modifiable risk factors—such as obesity, physical inactivity, poor dietary habits, tobacco use, and harmful alcohol consumption—as well as social determinants like income, education, and geographic region [11, 12]. In this context, individuals often present with more than one NCD (the co-occurrence of two or more chronic conditions in an individual), a phenomenon known as multi-morbidity [13].

Multi-morbidity has profound implications for patients and health systems alike. It complicates clinical management, increases healthcare costs, and heightens the risk of adverse outcomes including hospital admissions, disability, and early death [14, 15]. Yet, despite its significance, epidemiological studies in Uganda and similar LMIC settings have primarily focused on individual diseases in isolation. Traditional analytic methods, such as separate logistic regression models, fail to capture the interdependencies between conditions and may underestimate the true burden of shared risk exposures [16, 17].

Advanced statistical techniques, such as copula models, offer a robust solution for jointly analyzing multiple outcomes while preserving the distinct characteristics of each [18]. Copulas are flexible functions that allow for the modeling of complex dependence structures between variables, including those with non-linear or non-elliptical relationships [19]. By separating the marginal distribution of each outcome from their joint dependence, copula models provide a more accurate and nuanced understanding of disease clustering. Among available copula types, the Gaussian copula is particularly appealing due to its tractability [20], interpretability, and ability to model symmetric dependence structures—making it well-suited for multivariate binary outcomes such as those considered in this study [19].

Despite the growing burden of non-communicable diseases (NCDs) in low- and middle-income countries (LMICs), including Uganda, most epidemiological studies have traditionally focused on individual conditions in isolation [21, 22]. As a result, the complex interplay and co-occurrence of NCDs—such as hypertension, diabetes, and cardiovascular disease (CVD)—remain poorly understood, particularly in sub-Saharan Africa [3, 23]. While multi-morbidity has been studied in high-income settings using various multivariate methods [24], few, if any, studies in Uganda have jointly modeled the distribution and interdependence of these three major conditions using advanced dependence modeling. To our knowledge, this is the first study to apply a copula-based statistical framework to jointly examine hypertension, diabetes, and CVD in a nationally representative sample of Ugandan adults. By integrating survey-weighted logistic regression, multiple imputation for missing data, and copula models to estimate joint dependence, this study provides a methodologically novel and contextually relevant approach to understanding multi-morbidity patterns in a resource-limited setting.

This study aims to investigate the co-occurrence and shared determinants of hypertension, diabetes, and CVD among Ugandan adults using the copula framework. Specifically, we apply marginal logistic regression models to estimate disease-specific associations with sociodemographic, behavioral, and clinical covariates [25], and then use copula models to capture the joint distribution of the three conditions. This approach enables a deeper understanding of how these NCDs co-occur and what factors are associated with their clustering [1].

Understanding the joint distribution of chronic diseases is critical for public health planning and resource allocation. Identifying populations at higher risk of multi-morbidity can inform the design of integrated screening, prevention, and management programs. In Uganda, where health services are often fragmented and vertically oriented, a shift toward person-centered, comprehensive NCD care is urgently needed. The findings from this study contribute to the growing body of evidence supporting such a shift and provide methodological guidance for future epidemiological studies of multi-morbidity in LMICs.

## Materials and methods

### Study Design and Data Source

This study is a secondary data analysis of the 2014 Uganda WHO STEPwise approach to Surveillance (STEPS) survey, a nationally representative, cross-sectional household survey conducted to assess non- communicable disease (NCD) risk factors. The WHO STEPS methodology provides standardized procedures for data collection, allowing for international comparability and national monitoring [26–28]. The survey covered Ugandan adults aged 18 to 69 years and collected self-reported and clinical data on behavioral and biological NCD risk factors, including tobacco use, alcohol consumption, physical activity, diet, blood pressure, body mass index, and biochemical markers.

### Study Population and Inclusion Criteria

All respondents aged between 18 and 69 years with complete data on blood pressure, fasting glucose, and cardiovascular disease diagnosis were eligible for inclusion. Participants with missing values on these key variables were excluded through case-wise deletion to ensure consistency in joint modeling. After cleaning, the final analytical sample consisted of 3,987 individuals.

### Ethical statement

We analyzed the 2014 Uganda STEPS NCD Risk Factors Survey data which is publicly available at https://extranet.who.int/ncdsmicrodata/index.php/catalog/633/. No ethical approval was needed as the data were de-identified and ethical approval was obtained in the past from Saint Francis Hospital, Nsambya Institutional Review Board in 2006 and renewed in 2013. The Uganda Ministry of Health NCDs Desk provided the dataset for this analysis following our request.

### Variables and Measurement

#### Variable Coding and Recoding

Most variables were derived directly from the Uganda STEPS survey dataset and coded following WHO guidelines. However, several variables required recoding to align with analytical objectives. For instance, the region variable was constructed from district-level or ethnicity codes, using administrative mappings consistent with Uganda Bureau of Statistics classifications. The mapping process was carefully cross-checked using official district-region lookups, and ambiguous or unclassified entries were flagged and reviewed.

To minimize misclassification, a two-step validation was conducted: first, automated mapping using standardized codes, and second, manual review of outliers or unmatched entries by two independent analysts. In cases of unresolved ambiguity, such entries were excluded from regional comparisons to preserve data integrity.

#### Outcome Variables

Three binary health outcomes were jointly modeled; Hypertension status, Diabetes status and Cardiovascular disease status (CVD).

Systolic and diastolic blood pressure were recorded up to three times per respondent. Mean values were computed based on the second and third measurements (if available). A participant was classified as having high blood pressure (hypertension) if the mean systolic blood pressure was ≥140 mmHg or the mean diastolic was ≥90 mmHg. A composite variable for hypertension status was then derived, coded as positive if the participant self-reported a prior diagnosis (told by a health professional) or met the measurement criteria.

Diabetes status was determined using both self-report and biomarker data. Participants were coded as having diabetes if they; reported a prior diagnosis by a health professional, or had a fasting blood glucose level >7.0 mmol/L.

A variable indicating whether a respondent had been diagnosed with any form of cardiovascular disease was retained and renamed to cvd_status. This binary variable captures whether the respondent had ever been told by a health professional that they had a heart attack, stroke, or other cardiovascular condition. The variable was labeled accordingly for interpretability during analysis.

#### Predictor Variables

Several sociodemographic variables were recoded into analytically meaningful categories. The continuous age variable was grouped into four age bands: 18–29, 30–44, 45–59, and 60–69 years. Region of residence was derived from ethnicity codes, with ethnic groups being mapped to Uganda’s major geographic regions: Central, Western, Eastern, and Northern. Education level was recoded into four categories: no formal education, primary education, secondary education (including both O-Level and A-Level), and tertiary or university education (including vocational and postgraduate training). Marital status was categorized into three groups: unmarried (never married), married or cohabiting, and separated, divorced, or widowed. Monthly income was transformed into a new variable measured in United States dollars and grouped into three brackets: less than $277, between $278 and $2,777, and $2,778 or more.

Anthropometric and clinical indicators were computed based on WHO guidelines. Body Mass Index (BMI) was calculated as weight in kilograms divided by the square of height in meters, and then categorized into underweight (BMI <18.5), normal weight (BMI 18.5–24.9), overweight (BMI 25.0–29.9), and obese (BMI ≥30). Total cholesterol was categorized into normal (<6.2 mmol/L) and high (≥6.2 mmol/L) to indicate hypercholesterolemia. All generated and recoded variables were labeled appropriately to improve the clarity of output tables and ensure consistency in interpretation during statistical modeling. Table 1 summarizes the sociodemographic and clinical characteristics of the 3,987 adult participants in the 2014 Uganda NCD STEPS survey after data cleaning. Women comprised a slight majority, representing 52% of the weighted sample, compared to 48% men. Most participants resided in the Western (37.3%) and Northern (25.8%) regions, with smaller proportions from the Central (18.3%), Eastern (17.5%), and Other/Refused (1.0%) regions.

**Table 1.** Descriptive Statistics of the Study Variables.

| Variable | Code | Level | Frequency | Weighted | Percent |
| --- | --- | --- | --- | --- | --- |
| Sex | 0 | Men | 1604 | 5852164 | 48 |
|  | 1 | Women | 2383 | 6338942 | 52 |
| Region of Residence | 0 | Western | 778 | 1994480 | 18.29 |
|  | 1 | Eastern | 658 | 1912890 | 17.54 |
|  | 2 | Northern | 835 | 2817510 | 25.84 |
|  | 3 | Other/Refused | 44 | 107602.1 | 0.99 |
|  | 4 | Central | 1236 | 4072092 | 37.34 |
| Educational Attainment | 0 | No formal education | 654 | 1816860 | 14.94 |
|  | 1 | Primary | 1626 | 5022019 | 41.31 |
|  | 2 | Secondary | 1304 | 4211337 | 34.64 |
|  | 3 | Tertiary/University | 387 | 1106976 | 9.11 |
| Marital Status | 0 | Married/Cohabiting | 2867 | 8685992 | 71.28 |
|  | 1 | Separated/Divorced/<br>Widowed | 489 | 1122097 | 9.21 |
|  | 2 | Unmarried | 627 | 2377137 | 19.51 |
| Age Group | 0 | 18-29 | 1616 | 5516593 | 45.25 |
|  | 1 | 30-44 | 1399 | 4044370 | 33.17 |
|  | 2 | 45-59 | 699 | 1677130 | 13.76 |
|  | 3 | 60-69 | 273 | 953013.2 | 7.82 |
| Current Alcohol Use | 0 | No | 2092 | 6311155 | 51.83 |
|  | 1 | Yes | 1890 | 5866361 | 48.17 |
| Current Tobacco Use | 0 | No | 3638 | 11011854 | 90.42 |
|  | 1 | Yes | 345 | 1167057 | 9.58 |
| Vigorous Activity at Work | 0 | No | 2337 | 7205109 | 58.96 |
|  | 1 | Yes | 1569 | 5014771 | 41.04 |
| Hypertension Status | 0 | No Hypertension | 1099 | 3477821 | 62.85 |
|  | 1 | Hypertension | 674 | 2055857 | 37.15 |
| Diabetes Status | 0 | No Diabetes | 89 | 271044.2 | 35.02 |
|  | 1 | Diabetes | 188 | 502917.2 | 64.98 |
| Cardiovascular Disease<br>(CVD) | 0 | No | 3364 | 11140410 | 91.17 |
|  | 1 | Yes | 343 | 1078746 | 8.83 |
| Total Blood Cholesterol | 0 | High | 63 | 153359.9 | 1.25 |
|  | 1 | Normal | 3652 | 12090838 | 98.75 |
| Body Mass Index (BMI) | 0 | Normal weight | 330 | 8344412 | 68.29 |
|  | 1 | Under weight | 2531 | 1157307 | 9.47 |
|  | 2 | Overweight | 592 | 1675417 | 13.71 |
|  | 3 | Obese | 453 | 1042743 | 8.53 |

In terms of educational attainment, 41.3% had completed primary education, 34.6% secondary, 9.1% tertiary or university, while 14.9% reported no formal schooling. Regarding marital status, 71.3% were currently married or cohabiting, 19.5% were unmarried, and 9.2% were either separated, divorced, or widowed.

The age distribution was skewed toward younger adults, with 45.3% aged 18–29 years, 33.2% aged 30– 44, 13.8% aged 45–59, and 7.8% in the oldest group, 60–69 years.

Behavioral risk factors were common: 48.2% of adults reported current alcohol use, and 9.6% were current tobacco users. Regarding physical activity, 41.0% reported engaging in vigorous work-related activity.

In terms of clinical risk factors, 68.3% had a normal body mass index, while 13.7% were overweight, 8.5% obese, and 9.5% underweight. Notably, 62.9% of participants were classified as hypertensive, while 37.1% were normotensive. For diabetes, 65.0% were diagnosed or met criteria for diabetes, and 35.0% were non-diabetic. A smaller but significant proportion, 8.8%, reported a history of cardiovascular disease.

Hypercholesterolemia was rare, with only 1.25% having elevated total cholesterol (≥6.2 mmol/L).

These patterns underscore the dual burden of behavioral and clinical risk factors across Uganda’s adult population, with notable variation by sex, region, and age. The high prevalence of hypertension and diabetes, alongside substantial behavioral risk exposure, reflects the urgent need for targeted public health interventions.

### Statistical analysis

#### Application of Survey Design and Weighting

The analysis appropriately incorporated the complex survey design of the Uganda NCD STEPS survey by specifying primary sampling units (psu), strata (stratum), and sampling weights (wstep3) in all models using the `svydesign` function. All inferential analyses were conducted using the `svyglm` function, which accounts for clustering, stratification, and unequal probabilities of selection, yielding population-representative estimates and robust standard errors.

#### Handling of Missing Data

To ensure the validity of our findings, we carefully evaluated the nature and extent of missing data prior to modeling. Little’s MCAR test indicated strong evidence against the assumption that data were missing completely at random (χ² = 2624, df = 145, *p* < 0.001), suggesting that reliance on complete-case analysis alone could introduce bias. Therefore, we adopted multiple imputation as the primary approach for addressing missingness.

Multiple imputation was performed using the Multivariate Imputation by Chained Equations (MICE) method, implemented via the mice package in R. All variables included in the main analysis models— both predictors and outcomes—were incorporated into the imputation model, along with auxiliary variables hypothesized to predict missingness. Ten imputed datasets were generated using predictive mean matching (PMM), a method that is well-suited for categorical and semi-continuous variables and preserves the original distribution of the data.

Each imputed dataset was analyzed separately using logistic regression models for the three binary outcomes: hypertension, diabetes, and cardiovascular disease (CVD). The models included the same covariates as those used in the complete-case analysis. The resulting estimates were then combined using Rubin’s rules to produce pooled odds ratios (ORs), 95% confidence intervals (CIs), and p-values. This approach accounts for both within-imputation and between-imputation variability, leading to more robust and efficient inference compared to single imputation or case-wise deletion. Complete-case results were retained for comparison as part of a sensitivity analysis.

#### Marginal Models

Survey-weighted logistic regression models were independently fitted for each of the three binary outcomes: hypertension, diabetes, and cardiovascular disease (CVD). Each model adjusted for a common set of covariates—age group, sex, body mass index (BMI) category, marital status, region of residence, alcohol use, tobacco use, high cholesterol, and vigorous physical activity—to control for potential confounding. All models accounted for the complex survey design, including primary sampling units, stratification, and sampling weights, using the svyglm() function from the survey package in R. The estimated linear predictors from each model were transformed using the standard normal cumulative distribution function (CDF) to produce probability-scale residuals, which serve as pseudo-observations for copula modeling. This ensures that the marginal distributions are correctly specified and enables flexible modeling of joint dependence structures.

#### Copula Framework

To model the joint dependence structure among hypertension, diabetes, and CVD, we applied a copula-based approach, which allows for the separate modeling of marginal distributions and their dependencies. Copulas offer a flexible framework to capture complex, potentially nonlinear relationships between outcomes, which may not be adequately represented in standard multivariate models.

Two copula models were evaluated:

i. Clayton Copula, which captures lower-tail dependence—emphasizing the tendency for joint occurrence of extreme low outcomes.
ii. Gaussian Copula, which assumes a symmetric dependence structure and is suited for modeling moderate association in both tails.

Copula models provide a flexible framework that separates the modeling of marginal distributions from the dependence structure, enabling the modeling of complex, nonlinear associations between variables that traditional multivariate regression techniques may fail to capture [19, 29]. The tri-variate Gaussian copula is defined as: [18].

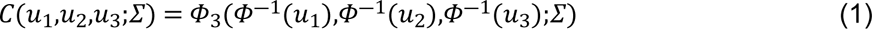

where:

- uᵢ = Fᵢ(Yᵢ) are the marginal CDFs of the binary outcomes Yᵢ,
- Φ⁻¹ is the inverse standard normal CDF,
- Φ₃ denotes the tri-variate standard normal CDF,
- and Σ is the correlation matrix governing the dependence structure among the three outcomes.

The parameters of the Gaussian copula, specifically the correlation coefficients in Σ, quantify the degree of association between each pair of outcomes on the probability scale. These parameters were estimated via maximum likelihood using pseudo-observations derived from the marginal models [30].

The trivariate Clayton copula is defined as:

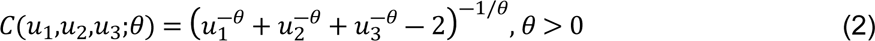

where:

- uᵢ = Fᵢ(Yᵢ) are the marginal CDFs of the binary outcomes Yᵢ,
- 𝜃 is the clayton copula parameter.

The Kendall’s tau for the Clayton copula is given by:

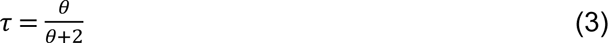

which provides a measure of the concordance between outcome pairs.

#### Software and Reproducibility

All analyses were conducted using R version 4.4.3 [31].The copula-based joint modeling of binary outcomes—hypertension, diabetes, and cardiovascular disease (CVD)—was implemented using the copula and `VineCopula` packages, which allowed for flexible modeling of dependence structures, including both Gaussian and Archimedean copulas. The `mvtnorm` package was used to handle multivariate normal distributions for marginal modeling. Data cleaning, transformation, and manipulation were conducted using `dplyr`, while `ggplot2` was used to visualize results and model diagnostics.[32], `forcats` for data manipulation and preprocessing [33]. The full R script used for data processing, model fitting, and output generation is available upon request to ensure reproducibility.

## Results

### Survey-Weighted Logistic Regression

#### Complete-Case Analysis (CCA)

Using survey-weighted logistic regression on complete cases, several associations were observed across the three binary outcomes (Table 2). Obesity was strongly associated with higher odds of hypertension (OR = 27.33; 95% CI: 2.77–269.6) compared to being underweight. Overweight also increased the odds of hypertension (OR = 8.51; 95% CI: 1.00–72.2) but was inversely associated with diabetes (OR = 0.17; 95% CI: 0.03–0.93). Vigorous physical activity at work was protective against hypertension (OR = 0.22; 95% CI: 0.09–0.56).

**Table 2.** Survey-Weighted Logistic Regression Results (Complete-Case Analysis) Odds ratios (OR) and 95% confidence intervals (CI) for predictors of hypertension, diabetes, and cardiovascular disease (CVD) using complete-case data.

| Variable | Variable | Hypertension OR (95% CI) | Diabetes OR (95% CI) | CVD OR (95% CI) |
| --- | --- | --- | --- | --- |
| Sex (Ref: Men) | Sex |  |  |  |
| Women | Women | 0.69 (0.27, 1.76) | 1.29 (0.36, 4.67) | 1.74 (0.62, 4.90) |
| Region (Ref: Central) | Region (Ref: Central) |  |  |  |
| Eastern | Eastern | 1.04 (0.37, 2.93) | 0.96 (0.28, 3.31) | **0.11 (0.01, 0.91)** |
| Northern | Northern | 0.63 (0.21, 1.87) | 1.32 (0.38, 4.61) | 0.75 (0.19, 2.96) |
| Other/Refused | Other/Refused | 3.90 (0.15, 99.7) | 2.62 (0.08, 86.6) | 2.00e-7 (0, NA) |
| Western | Western | 0.92 (0.34, 2.50) | 0.82 (0.25, 2.71) | 0.32 (0.07, 1.48) |
| Marital Status (Ref: Married) | Marital Status (Ref: Married) |  |  |  |
| Separated/Divorced/Widowed | Separated/Divorced/Widowed | 4.58 (0.99, 21.2) | 1.55 (0.32, 7.58) | 1.12 (0.32, 3.97) |
| Unmarried | Unmarried | 0.46 (0.12, 1.73) | 0.84 (0.17, 4.18) | 0.35 (0.06, 2.07) |
| BMI Category(Ref: Underweight) | BMI Category (Ref: Underweight) |  |  |  |
| Normal weight | Normal weight | 4.11 (0.53, 31.8) | 0.60 (0.10, 3.48) | 1.18 (0.18, 7.70) |
| Overweight | Overweight | **8.51 (1.00, 72.2)** | **0.17 (0.03, 0.93)** | 0.42 (0.04, 4.56) |
| Obese | Obese | **27.33 (2.77, 269.6)** | 0.64 (0.09, 4.80) | 4.01 (0.37, 43.1) |
| Age Group (Ref: 18–29) | Age Group (Ref: 18–29) |  |  |  |
| 30–44 | 30–44 | 2.46 (0.79, 7.66) | 1.46 (0.38, 5.57) | 4.18 (0.93, 18.8) |
| 45–59 | 45–59 | **4.14 (1.28, 13.4)** | 2.50 (0.62, 10.1) | **7.54 (1.58, 36.1)** |
| 60–69 | 60–69 | 4.18 (0.86, 20.3) | 1.22 (0.18, 8.38) | 2.68 (0.36, 20.2) |
| Alcohol Use (Ref: No) | Alcohol Use |  |  |  |
| Yes | Yes | 0.55 (0.25, 1.22) | **0.35 (0.14, 0.90)** | 1.89 (0.62, 5.81) |
| Tobacco Use (Ref: No) | Tobacco Use |  |  |  |
| Yes | Yes | 2.72 (0.21, 35.3) | 3.23 (0.38, 27.6) | 2.64 (0.22, 31.2) |
| High Cholesterol (Ref: High) | High Cholesterol |  |  |  |
| Normal | Normal | 2.09e-7 (0, 26.1) | 2.28 (0.40, 13.1) | 4.22 (0.56, 31.7) |
| Vigorous Activity (Ref: No) | Vigorous Activity |  |  |  |
| Yes | Yes | **0.22 (0.09, 0.56)** | 0.66 (0.23, 1.92) | 1.29 (0.32, 5.25) |

**\*\* Bolded estimates** indicate statistical significance (95% CI does not cross 1.0), Ref = Reference category & OR = Odds Ratio; CI = Confidence Interval.

Older age groups had elevated odds of CVD. Specifically, respondents aged 45–59 years had over seven times the odds of CVD compared to the 18–29 age group (OR = 7.54; 95% CI: 1.58–36.1). Regionally, residence in the Eastern region was associated with lower odds of CVD (OR = 0.11; 95% CI: 0.01–0.91) relative to the Central region.

#### Multiple Imputation Analysis with Survey Weights

Results from the multiply imputed, survey-weighted models (Table 3) were generally consistent with the CCA but showed increased statistical efficiency and precision. Women had lower odds of hypertension (OR = 0.48; 95% CI: 0.32–0.72) but higher odds of CVD (OR = 1.86; 95% CI: 1.23–2.47) compared to men. Obesity remained a significant predictor of hypertension (OR = 2.00; 95% CI: 1.02–3.92), and higher BMI was associated with substantially lower odds of diabetes. For instance, those categorized as overweight had 91% lower odds of diabetes (OR = 0.09; 95% CI: 0.03–0.28).

**Table 3.**
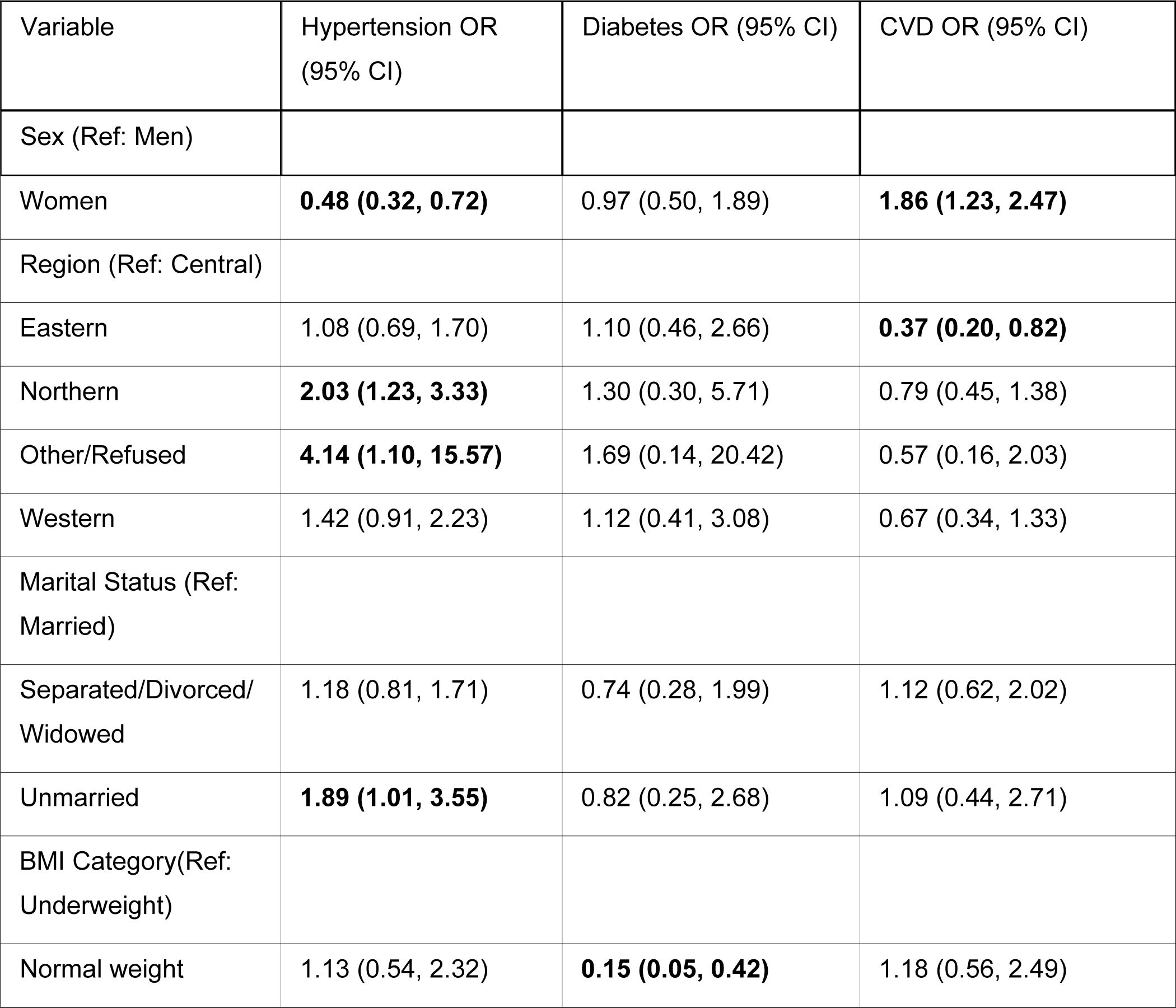
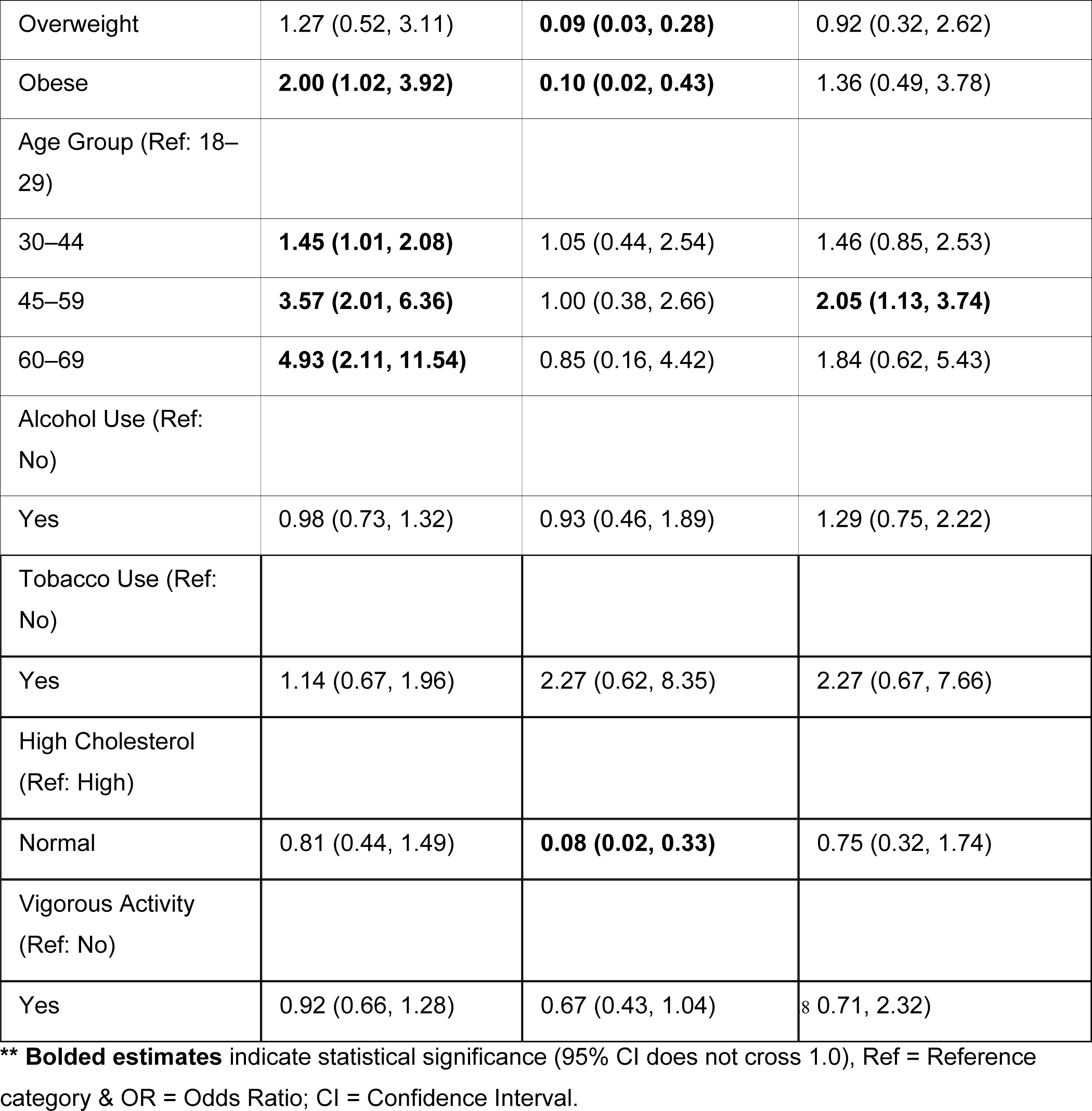
Survey-Weighted Logistic Regression Results (Multiple Imputation) Odds ratios (OR) and 95% confidence intervals (CI) for predictors of hypertension, diabetes, and CVD after multiple imputation.

Age-related trends persisted: participants aged 60–69 had nearly fivefold increased odds of hypertension (OR = 4.93; 95% CI: 2.11–11.54) and elevated CVD odds. Living in the Eastern region was associated with decreased odds of CVD (OR = 0.37; 95% CI: 0.20–0.82), while the Northern region was linked to higher odds of hypertension (OR = 2.03; 95% CI: 1.23–3.33). Normal cholesterol levels strongly protected against diabetes (OR = 0.08; 95% CI: 0.02–0.33).

### Dependence Structure Among NCD Outcomes

To evaluate the joint dependence among hypertension, diabetes, and cardiovascular disease (CVD), we fitted two copula models using probability-scale residuals from survey-weighted logistic regressions.

The Clayton copula estimated a dependence parameter (θ) of 0.0757, corresponding to a low Kendall’s τ of 0.0365, indicating minimal lower-tail dependence between the NCD outcomes. The log-likelihood for this model was 87.33, and the Akaike Information Criterion (AIC) was −172.65.

The Gaussian copula, which captures symmetric dependence, showed a stronger fit with a log-likelihood of 184.04 and a lower AIC of −362.08, indicating a substantially better model. The estimated dependence parameters (correlation coefficients) for the Gaussian copula were −0.025, −0.122, and 0.377, reflecting weak to moderate pairwise associations.

Overall, the Gaussian copula outperformed the Clayton copula in capturing the dependence structure among the NCD outcomes, suggesting that a symmetric, moderate joint dependency better characterizes multi-morbidity patterns in this population.

### Diagnostic Visualization

Visual inspection of the probability-scale residuals (Figure 1) confirmed reasonable alignment with the assumptions of the copula model. Overall, the Gaussian copula provided a flexible framework to jointly model the marginal distributions of the three disease outcomes while capturing their underlying correlation.

**Fig 1.**
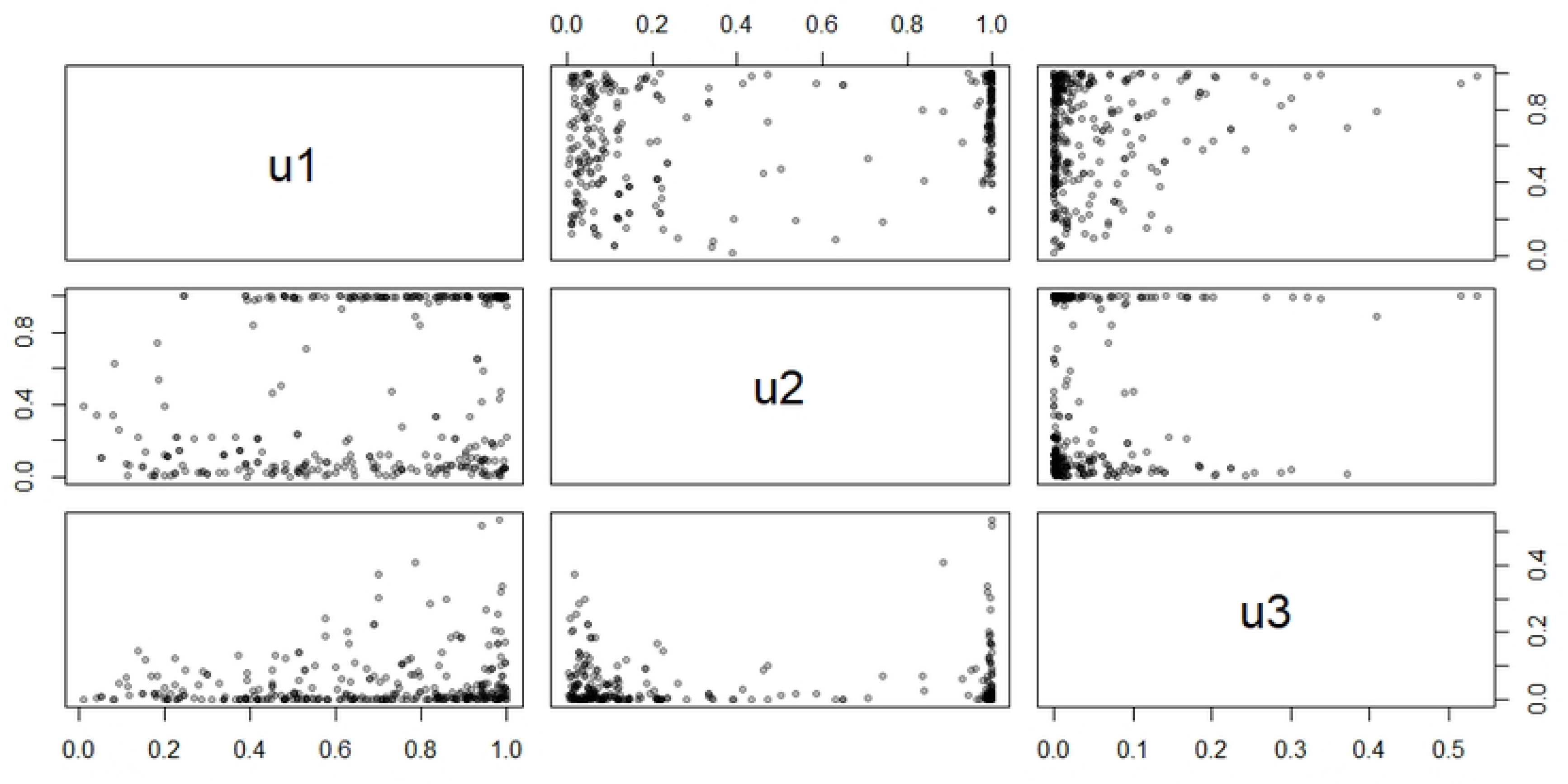
Visual inspection of the probability-scale residuals.

## Discussion

This study provides a comprehensive epidemiological analysis of three major non-communicable diseases (NCDs)—hypertension, diabetes, and cardiovascular disease (CVD)—using nationally representative data from the 2014 Uganda NCD STEPS survey [1, 2]. By integrating survey-weighted logistic regression models with multiple imputation for missing data and copula-based dependence modeling, we provide a nuanced understanding of both individual disease risk factors and their interrelationships within the Ugandan population [34–37].

### Key Findings

Our results reaffirm well-established determinants of NCDs. Increasing age and higher body mass index (BMI) were strongly associated with higher odds of hypertension and CVD [13, 25]. Specifically, individuals aged 45–59 and 60–69 exhibited significantly greater odds of hypertension and CVD compared to those aged 18–29 [38]. Similarly, obesity was associated with more than twice the odds of hypertension and modestly higher odds of CVD [9, 14, 15]. These associations reflect ongoing demographic and nutritional transitions in sub-Saharan Africa, where urbanization, sedentary lifestyles, and dietary shifts have contributed to rising NCD prevalence [25, 38, 39].

Interestingly, diabetes showed a more complex pattern. In the complete-case analysis, overweight and obese individuals were less likely to report diabetes, which contradicts conventional metabolic risk profiles. However, this inverse association diminished in the multiple imputation models, suggesting the possibility of selection bias or underreporting among specific subpopulations [10, 15–17]. It may also reflect the limitations of self-reported diagnosis, particularly in low-resource settings where access to diagnostic services is limited.

Sex-specific patterns were also observed. While female sex was not consistently associated with diabetes or CVD, it was significantly protective against hypertension in the multiple imputation analysis [12]. This aligns with findings from other African studies that highlight lower hypertension prevalence among women, potentially due to hormonal, behavioral, or healthcare utilization differences [4–6]. Regionally, individuals residing in the Northern and Other/Refused regions exhibited higher odds of hypertension but lower odds of CVD compared to those in Central Uganda, underscoring regional disparities in disease burden and health access [12, 40–42].

Lifestyle factors such as alcohol and tobacco use were not consistently associated with NCD outcomes, which may reflect limited statistical power, reporting biases, or heterogeneous patterns of use. However, vigorous physical activity at work was inversely associated with hypertension in the complete-case model, supporting the role of occupational activity in cardiovascular health [40–42].

### Copula-Based Dependence Modeling

A major innovation of this study is the use of copula models to explore the dependence structure between hypertension, diabetes, and CVD [18–20, 29, 30, 43, 44]. While marginal models identify disease-specific risk factors, copulas allow for the joint modeling of multiple outcomes, providing insight into their co-occurrence and shared etiology.

We found that the Gaussian copula offered a significantly better fit than the Clayton copula, as evidenced by a higher log-likelihood and lower AIC. The estimated correlation coefficients from the Gaussian copula indicated moderate positive associations between disease pairs—particularly between hypertension and CVD—consistent with their overlapping pathophysiological pathways (e.g., endothelial dysfunction, atherosclerosis). The Clayton copula, which captures asymmetric lower-tail dependence, yielded a much lower Kendall’s tau (τ = 0.0365), suggesting minimal left-tail association. This further supports the suitability of the symmetric Gaussian structure for modeling joint NCD outcomes in this population.

These findings emphasize that although hypertension, diabetes, and CVD share common risk factors, they do not cluster strongly in all individuals [19, 45–47]. The moderate dependence observed suggests partial overlap in their occurrence, likely influenced by individual-level behaviors, genetic predispositions, and differential access to care.

### Strengths and Limitations

This study has several strengths. First, it applies complex survey-weighted regression techniques, accounting for stratification, clustering, and unequal probabilities of selection, thereby ensuring population-level generalizability. Second, it addresses missing data through multiple imputation, reducing potential bias from complete-case analysis. Third, the use of copula models represents an advanced methodological approach that extends beyond conventional regression, offering a flexible and robust framework for multivariate disease analysis.

However, some limitations must be acknowledged. The cross-sectional design of the STEPS survey limits causal inference. Additionally, reliance on self-reported data for some conditions (e.g., CVD) may introduce misclassification bias. Moreover, the copula approach assumes correctly specified marginal models; misspecification at this stage could bias dependence parameter estimates.

The definitions used for hypertension and diabetes in this study combined self-reported diagnosis with biomarker thresholds, aligning with WHO STEPS survey guidelines and enhancing the sensitivity of case detection. However, this approach may introduce differential misclassification, particularly for diabetes. Individuals aware of their diagnosis (and possibly under treatment) may meet only the self-reported criterion without exhibiting elevated blood glucose at the time of the survey. Conversely, some individuals with undiagnosed hyperglycemia may have been missed if biomarker data were missing or imprecise. These dynamics could lead to either overestimation or underestimation of true prevalence and may affect the precision of estimated associations.

The classification of cardiovascular disease (CVD) relied solely on self-reported prior diagnosis by a healthcare professional. This method is subject to significant limitations in low- and middle-income country (LMIC) settings like Uganda, where under-diagnosis and limited access to cardiovascular diagnostic services are well-documented. As a result, the prevalence of CVD reported here (approximately 9%) likely underrepresents the true burden. This under-ascertainment introduces the risk of outcome misclassification bias—particularly non-differential misclassification, which tends to attenuate associations toward the null. Consequently, some true associations with CVD risk factors may have been underestimated in our analyses.

Future surveys would benefit from incorporating objective cardiovascular assessments such as electrocardiograms or validated symptom checklists to improve case detection. In the meantime, readers should interpret the findings related to CVD with caution, acknowledging that the observed estimates likely reflect a conservative approximation of the true prevalence and associations.

Finally, due to convergence issues, we were unable to apply weights directly within the copula framework, and thus, the dependence estimates may not fully reflect population structure.

### Public Health and Research Implications

The observed interdependence among hypertension, diabetes, and CVD supports the growing call for integrated NCD prevention and control strategies in low- and middle-income countries (LMICs). Rather than treating these conditions in isolation, primary healthcare systems should adopt a syndemic approach—recognizing the shared risk environment and reinforcing joint screening, education, and management.

Regionally targeted interventions are also warranted, particularly in areas with elevated risk. Health promotion efforts should prioritize obesity prevention and the early detection of hypertension, especially among older adults and those engaged in sedentary occupations. Further research is needed to validate these findings longitudinally and to explore the role of emerging risk factors such as stress, diet, and air pollution in the Ugandan context.

## Conclusion

This study provides a robust, population-level analysis of the prevalence and interrelationship of hypertension, diabetes, and cardiovascular disease in Uganda using nationally representative data. By combining survey-weighted logistic regression, multiple imputation, and advanced copula modeling, we offer a comprehensive view of both independent and joint disease risk. Our findings confirm known associations with age, BMI, and region, while also highlighting modest but non-negligible dependence between these NCDs—particularly between hypertension and CVD.

The superior fit of the Gaussian copula over the Clayton copula suggests that symmetric associations are a better representation of the joint burden of these conditions. These insights emphasize the importance of integrated NCD screening and management strategies in Uganda’s health system and underscore the need for regionally tailored public health interventions. Future longitudinal studies are warranted to explore causal pathways and to validate the joint modeling approach in diverse LMIC settings.

## Data Availability

The data underlying the results presented in the study are available from https://ghdx.healthdata.org/record/uganda-stepsnoncommunicable-disease-risk-factors-survey2014.

## Acknowledgments

The authors would like to thank the Uganda Ministry of Health, Non-Communicable Diseases (NCDs) Desk for unconditionally supporting this study by granting data access and providing administrative clearance.

